# A unified general cognitive factor explains impairment across schizophrenia, bipolar disorder, OCD and substance use disorder

**DOI:** 10.64898/2025.12.24.25342965

**Authors:** Pavithra Jayasankar, Srinivas Balachander, Swarna Buddha Nayok, Arushi Singh, Pavithra Dayalamurthy, Mino Susan Joseph, Aswathy Das, Mahashweta Bhattacharya, Alka Shaktan, Chandrakanta S Hiremath, Aishwarya L Solanki, Gopika Jayaraj, Mallappagiri Sreenivasalu, N Shruthi, A R Alagarsami, Farooq Ali Syed, Pratibha Vinod, Vasundhra Teotia, Aina Kurian, J V Nikhilan, K Rajkumar, Chidatma Arampady, Tallapally Manideepika, Shama Shareen, P T Arsharani, V Manjula, Dhruva Ithal, Jayant Mahadevan, Ravi Kumar Nadella, Vanteemar S Sreeraj, Ganesan Venkatasubramanian, John P. John, Vivek Benegal, YC Janardhan Reddy, Mathew Varghese, Sanjeev Jain, Urvakhsh Meherwan Mehta, ADBS-CBM Consortium, Bharath Holla, Biju Viswanath

## Abstract

**Background:** Emerging evidence suggests that cognitive impairment cuts across traditional psychiatric diagnoses and may reflect a shared underlying cognitive liability. We examined whether a general cognitive factor (gFc) accounts for transdiagnostic deficits across schizophrenia (SZ), bipolar disorder (BD), obsessive-compulsive disorder (OCD) and substance use disorder (SUD).

**Methods:** A total of 472 affected individuals and 253 population healthy controls (HC) completed a standardized cognitive battery. Bifactor confirmatory factor analysis was used to derive general cognitive factor (gFc) and domain-specific abilities(cognitive flexibility, working memory, new learning and social cognition). Model fit supported the bifactor structure. Linear mixed-effect models, with family as random effect, and demographic variables as fixed effect evaluated transdiagnostic and diagnosis specific impairments..

**Results:** The gFc was significantly reduced across all diagnoses groups relative to HC, [B(SE): BD −0.49 (0.07), OCD −0.21 (0.07), SUD −0.43(0.08), SZ −0.76 (0.08), all p < 0.001]; verbal learning showed additional diagnosis-specific deficits, most pronounced in SZ [B (SE): −0.59 (0.09)] and BD[B(SE): −0.48 (0.09)], both p <0.001.

**Conclusion:** A single gFc accounts for substantial cognitive dysfunction across major psychiatric disorders, while verbal-learning impairment appears diagnosis-specific. These findings support a dimensional architecture of cognition in psychiatry and identify gFc as a promising transdiagnostic target for clinical and mechanistic research.

## Introduction

Major psychiatric illnesses (MPIs) are among the leading contributors to global disability and disease burden, significantly impairing social and occupational functioning(1). Despite decades of research, understanding their neurobiological underpinnings remains challenging, partly because traditional diagnostic categories describe complex behavioural syndromes rather than the underlying disease processes(2,3). These categories often obscure substantial within-disorder heterogeneity and the high prevalence of psychiatric comorbidity(4). Moreover, familial aggregation patterns indicate that the heritability is only partially specific to individual diagnoses(5,6). Growing evidence suggest that MPIs share phenotypic and biological features, challenging the assumption that they are diagnostically separate entities (7,8).

Emerging genetic and epidemiological studies have highlighted overlapping vulnerability mechanism across MPIs. Large-scale genomic analyses show substantial shared genetic architecture among psychiatric disorders, suggesting the presence of transdiagnostic risk factors(8,9).This has led to conceptualisation of dimensional approaches, which aim to capture shared liability across disorders and disorder-specific variance. One such conceptual framework is the general psychopathology factor (“*p factor”*) proposed by Caspi and colleagues reflecting a single latent dimension of liability that spans multiple psychiatric conditions(10).

Cognitive impairment is a hallmark of MPIs and increasingly recognised as a promising transdiagnostic marker(11–13). Deficits in domains such as working memory, processing speed, new learning and social cognition have been consistently reported across psychiatric disorders (14–16). Based on this evidence, Abramovitch and colleagues proposed a general cognitive factor (“C factor”), representing a latent construct that captures shared cognitive deficits across disorders (12). While prior studies often rely on meta-analytic syntheses or heterogenous datasets, the generalisability of these findings is limited, highlighting the need for single-cohort studies that assess cognition across multiple MPIs using unform measures.

In addition to examining transdiagnostic cognitive deficits, it is important to investigate how these deficits relate to clinical severity and treatment. Cognitive functioning may be influenced by illness severity, duration as well as pharmacological interventions(17–19). Evaluating the relationship between latent cognitive factors and clinical measures can provide a more nuanced understanding of the determinants of cognitive deficits across MPIs.

To address these questions, we examined cognitive functioning in a large cohort of individuals affected with MPIs (SZ, BD, OCD and SUD), recruited from a longitudinal family based cohort study “Accelerated Discovery of Bran Disorder using Stem cells – Centre for Brain and Mind (ADBS-CBM)(20). Using both raw neuropsychological test scores and latent cognitive constructs representing specific domains and a general cognition factor, we aimed to characterize transdiagnostic and disorder-specific impairment. Using linear-mixed effects modelling to account for demographic variation and within-family clustering, we assess differences in general cognitive factor and specific domains across four diagnostic groups compared with healthy controls (HC). We hypothesize that 1) a general cognitive factor deficit will be evident across disorders, consistent with the C-factor model, and 2) specific diagnoses groups may show more pronounced deficits in certain domains, reflecting unique illness characteristics.

Furthermore, we explored associations between cognitive deficits, clinical severity and treatment variables providing a comprehensive assessment of cognitive dysfunction across MPIs.

## Methods

### Subjects

The study sample included patients diagnosed with MPI according to the DSM-IV criteria, namely SZ (n= 95 ), BD (n = 120), OCD (n = 124) and SUD (n = 133), multiple diagnosis (more than 1 of the above 4 diagnoses) (n = 72), and HC (n = 253). All the affected individuals were recruited from the adult psychiatry services and specialty psychiatry clinics of the National Institute of Mental Health & Neurosciences (NIMHANS), Bengaluru, India. The HC were recruited from the community by word-of- mouth, local advertisements, and from a previous study conducted at the same centre with similar methodology(21). All psychiatric diagnoses were corroborated by 2 trained psychiatrists. All the participants were screened using M.I.N.I till March, 2020 and FLI-11 (22) was added to the protocol later. The HC did not have any Axis-1 psychiatric diagnosis (as per M.I.N.I/FLII-11) and had no family history of MPI in their first-degree relatives. The study protocol and recruitment procedure are detailed in earlier publications(20,23) .

The NIMHANS ethics committee approved the study protocol and written informed consent was obtained from all the participants in the study.

### Neuropsychological assessments

Cognitive assessments were administered by trained clinical psychologists in a quiet setting during a single 40 to 50 minutes session. The order of the tests were counterbalanced. The following 4 tests were chosen based on the existing literature indicating varying levels of deficits in the 4 disorders under study and importantly, these tests were less sensitive to the influences of language and cultural bias.

#### Color Trails (CT) Test A and B(24)

Assessed processing speed and cognitive flexibility. Total time, errors and effective time were calculated. An “effective time-per-circle” (ETC)measure was calculated using the formula [TT/CC + TE/TC], where TT is the total time taken, CC is number of circles completed, TE is total errors committed and TC is total circles in the test (e.g., 25 for CT-A and 50 for CT-B(25).Scores were reversed so that higher values indicated better performance.

#### *The Auditory N-Back* (1-Back and 2-Back)

Assessed verbal working memory(26) . Thirty randomly ordered consonants common in Indian languages were presented (27). Total number of commissions (hits) and omissions(errors) were recorded. The outcome measure was accuracy which was calculated using the formula [1 – [number of commission errors + number of omission errors)/total trials ×100].

#### Rey’s Auditory Verbal Learning Test (RAVLT)

Measured verbal learning and memory(28). The first set of 15 words of familiar objects (list A), was presented over 5 trials and recall was done after trial. Subsequent to the 5^th^ trial, a separate interference trial of 15 objects (list B) was presented and recall was elicited for list A. The words have been previously translated to Indian languages Hindi, Kannada, Tamil, Telugu and Malayalam (27). The outcome measures were RAVLT total score (total words correctly recalled over 5 trials) and RAVLT immediate recall (total words correctly recalled after the interference trial).

#### Social Cognition Rating Tools in Indian Setting (SOCRATIS)

Measured social cognition in Indian sociocultural settings. Subsets from SOCRATIS were used to assess the second-order theory of mind since this measure showed consistent associations with real-world outcomes (29) in earlier studies. Adapted versions of 2 false belief stories – Ice-cream man task (30) and hidden banana task (31) and 2 irony detection stories (32) were administered. A second order theory of mind index (33) was calculated as the proportion of correct responses on these questions after accounting for responses to control questions.

### Clinical and Treatment variables

Illness severity and chronicity were measured using Clinical Global Impression-Severity(CGI-S)(34) and duration of illness. The treatment variables included the presence or absence of antipsychotics, mood stabilisers, anti-depressants and benzodiazepines (Binary coded, 0 = No, 1 = Yes).

### Statistical analysis

Descriptives statistics were computed using means (SD) for continuous variables and counts (%) for categorical variables. Group differences were compared using ANOVA for continuous variables and chi-square tests for categorical variables.

Missing cognitive data were imputed when ≤ 3 out of 7 test variables were missing per participant using multivariate imputation by chained equation (MICE) with a random forest-based algorithm(35). Among 797 total participants, 89.80% did not have any missing data.

A bifactor confirmatory factor analysis (BICFA) was performed to extract the latent cognitive domains - cognitive flexibility (CF), working memory (WM), new learning (NLR) and Social Cognition (SC) and a higher-order general cognitive factor (gFc). This was chosen as we aimed to validate a predefined factor structure rather than explore unknown patterns in the data (36). This approach captures variances that are shared, as well as unique to each cognitive domain, while addressing multidimensionality and enhancing the interpretability of our findings (36). The model fit was assessed using several goodness-of-fit indices. The model fit was considered good if the comparative fit index (CFI) and Tucker-Lewis’s index (TLI) were > 0.95 and the root mean squared error of approximation (RMSEA) < 0.06(37). The factor scores were then used as outcome (dependent) variables for all subsequent analyses.

Separate linear mixed-effects models were applied for raw test scores and latent factors, with group as a fixed effect, age, sex, and years of education as covariates, and family ID as a random effect. Spearman correlations were conducted to explore associations of cognitive performance with illness duration and severity respectively.

To explore the association between psychotropic use and cognitive performance, we fitted separate linear mixed-effects model for each latent factor with 4 treatment variables entered simultaneously as fixed-effect predictors along with age, sex, years of education and diagnosis.

Because multiple linear mixed-effects models were estimated across cognitive domains and diagnostic groups, p-values for all fixed-effect estimates were corrected using the False Discovery Rate (FDR) method (38). An FDR-adjusted p-value < 0.05 was considered statistically significant.

All analyses were performed in the R environment for statistical computing, version 4.4.1, using base packages, mice (39) , lavaan (40) and lme4(41).

## Results

### Sample Description

Table 1 shows the demographic and clinical characteristics of the individuals (n = 797) across 7 groups: SZ, BD, OCD, SUD, Multiple diagnosis, and HC. Of the 72 individuals classified under multiple diagnoses, 69 had either SZ or BD as their primary diagnoses.

**Table 1.**
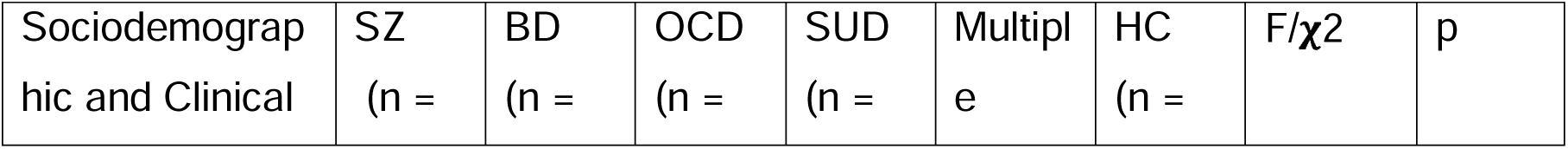
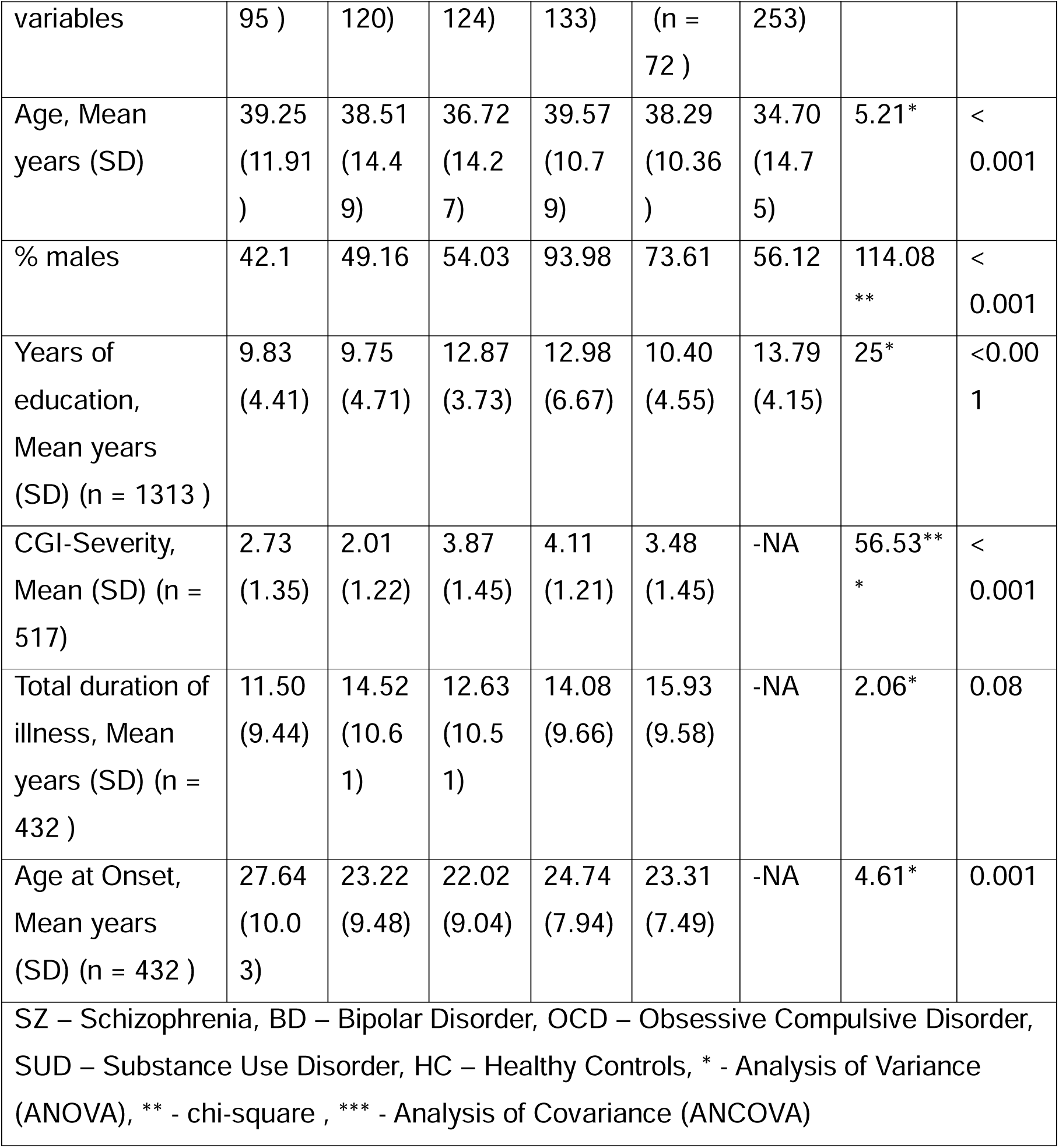
Sociodemographic and Clinical description of the study sample (n = 797)

Significant differences (p < 0.001) were observed in mean age, years of education, and CGI-severity across groups. The mean CGI-S was highest for SUD followed by SZ. The total illness duration showed no significant differences.

### Analysis of performance across cognitive tests (raw scores)

Significant lower scores (p < 0.007) in N-Back 1 , N-Back 2, AVLT (total & recall) were noted in SZ, BD, SUD and multiple diagnosis, in comparison with HC. Individuals with SZ had impairment in Color Trails 2 and the second-order theory of mind. The following associations (lower scores) were found significant at p<0.05: BD on Color Trails 2; OCD on N-Back 2 and AVLT (Total); and SZ on Color Trails 1 (Supplement table S2).

### Confirmatory Factor Analysis

Figure 2 shows the bifactor confirmatory factor analysis model. The model fit indices were CFI = 0.98, TLI = 0.96, and RMSEA = 0.05. These indicate that the model fit was excellent (42).

**Figure 2.**
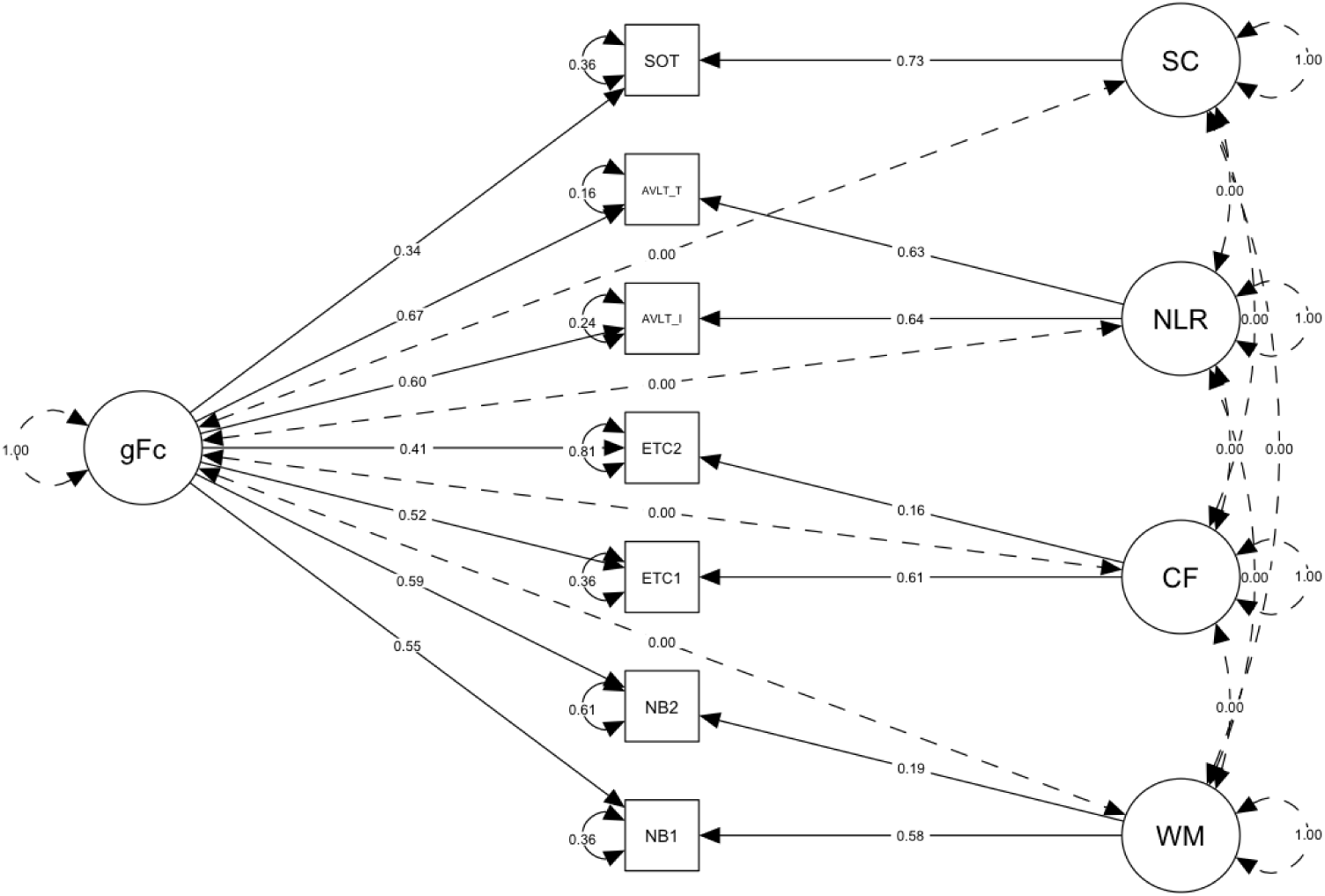
Bifactor Confirmatory Factor analysis (BICFA)

### Analysis of latent cognitive domains

Global cognitive ability (gFc) domain was significantly lower across all diagnoses (Figure 3). SZ, BD, and multiple diagnosis groups showed diagnosis-specific deficits in NLR. There were no statistically significant differences in CF, WM and SC in any of the studied disorders. The full results for the statistical models are shown in **Table 2**.

**Figure 3.**
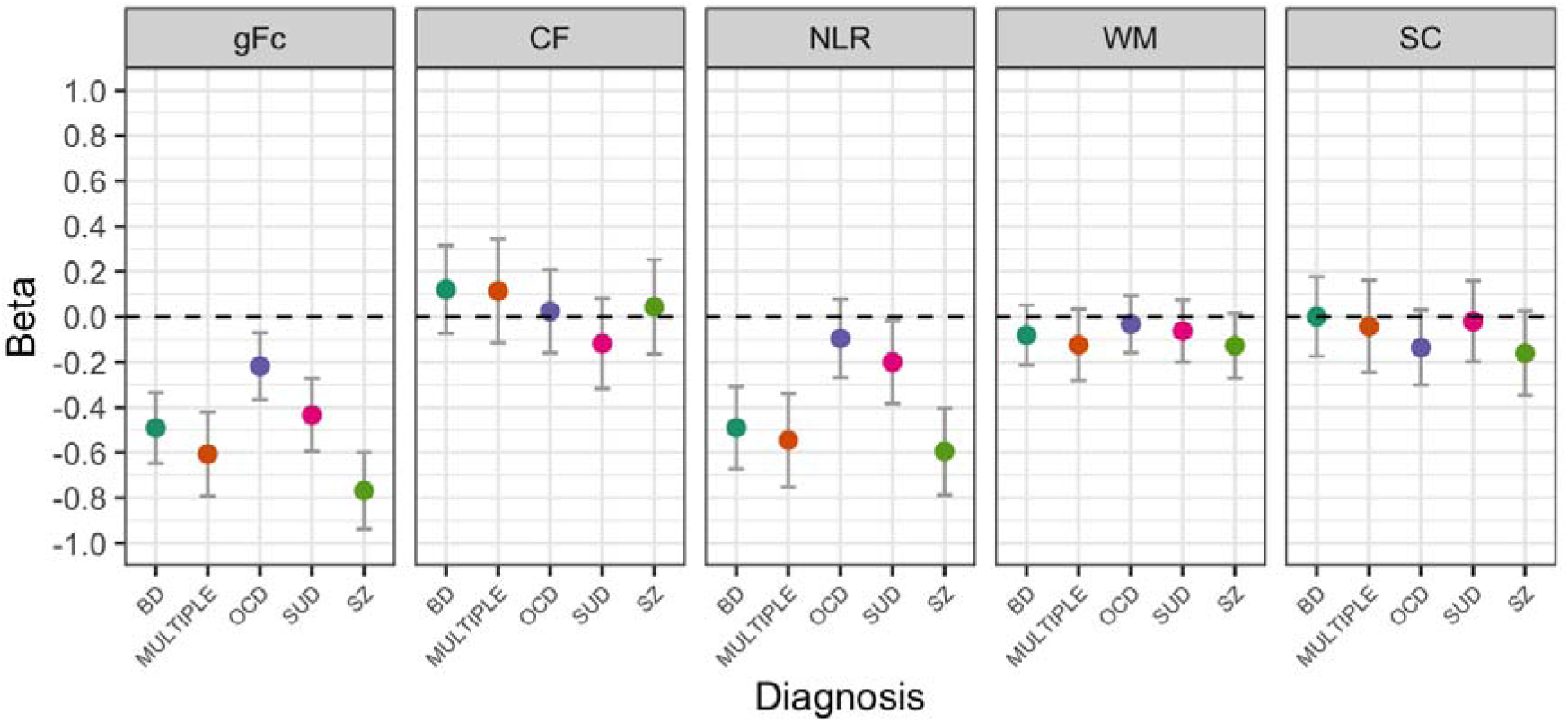
Standardized beta coefficients and standard error (+/- 2) for the neurocognitive domains among Major Psychiatric Illness (MPI) in comparison with population healthy control (PHC)(controlled for age, sex, years of education and family ID)

**Table 2.** Effect of diagnosis on latent domains of neurocognition using linear mixed effects analysis covarying for age, gender and years of education * p < 0.05, ** p < 0.01.

| Fixed effects | gFc | NLR | WM | CF | SC |
| --- | --- | --- | --- | --- | --- |
| Predictors of Interest: Diagnoses |  |  |  |  |  |
| BD | -0.49<br>[-0.65, -0.33]** | -0.48<br>[-0.67, -0.31]** | -0.08<br>[-0.21, 0.05] | 0.12<br>[-0.07, 0.31] | 0.0007<br>[-0.17, 0.18] |
| Multiple diagnosis | -0.60<br>[-0.79, -0.42]** | -0.54<br>[-0.75, -0.34]** | -0.12<br>[-0.28, 0.03] | 0.11<br>[-0.12, 0.34] | -0.04<br>[-0.25, 0.16] |
| OCD | -0.21<br>[-0.37, -0.07]** | -0.09<br>[-0.27, 0.08] | -0.03<br>[-0.16, 0.10] | 0.02<br>[-0.16, 0.21] | -0.13<br>[-0.30, 0.03] |
| SUD | -0.43<br>[-0.60, -0.27]** | -0.20<br>[-0.38, -0.02]* | -0.06<br>[-0.20, 0.08] | -0.11<br>[-0.32, 0.08] | -0.01<br>[-0.20, 0.16] |
| SZ | -0.76<br>[-0.94, -0.60]** | -0.59<br>[-0.79, -0.40]** | -0.12<br>[-0.27, 0.02] | 0.04<br>[-0.16, 0.25] | -0.16<br>[-0.35, 0.03] |
| Covariates |  |  |  |  |  |
| Age | -0.01<br>[-0.02, -0.01]** | -0.01<br>[-0.02, -0.01]** | 0.001<br>[-0.00, 0.00] | -0.001<br>[-0.01, 0.00] | -0.002<br>[-0.00, 0.00] |
| Gender (Male) | 0.02<br>[-0.08, 0.13] | -0.34<br>[-0.46, -0.23]** | 0.16<br>[0.08, 0.26]** | 0.14<br>[0.02, 0.28]* | -0.14<br>[-0.26, -0.03] |
| Years of education | 0.07<br>[0.06, 0.08]** | 0.01<br>[0.00, 0.03]* | 0.013<br>[0.00, 0.02]** | 0.02<br>[0.01, 0.04]** | 0.014<br>[0.00, 0.03] |

BD – Bipolar disorder, OCD – Obsessive compulsive disorder, SUD – substance use disorder, SZ – schizophrenia, gFc – global neurocognition, NLR – new learning and recall, WM – working memory, CF – cognitive flexibility, SC – social cognition

### Association with illness chronicity

Total duration of illness showed a small but significantly negative correlation with gFc (r = −0.16, p <0.001) and NLR (r = −0.18, p <0.001). No significant correlations were observed with TDI and other latent cognitive domains (Supplementary table S3).

### Association with illness severity

There were no significant correlations between CGI-S and any of the latent cognitive domains including gFc, NLR, WM, CF and SC (Supplementary table S3).

### Association with treatment variables

Antipsychotic use was not associated with gFc or any specific cognitive domain, except for lower social cognition scores at trend-level significance (did not survive FDR corrections) (β = –0.25, 95% CI: –0.46 to –0.03). Mood stabiliser use did not show significant associations with any of the cognitive domains. Antidepressant use was associated with slightly better gFc (β = 0.24, 95% CI: 0.02 to 0.45), but was not associated with NLR, CF, WM and SC. Benzodiazepine use did not show statistically significant associations with any latent cognitive domain. Full model estimates are provided in Supplementary Table S4.

## Discussion

Our study examined transdiagnostic and disorder-specific cognitive impairment across MPI within a single, well-characterized cohort using both raw neuropsychological test scores and latent cognitive constructs. Consistent with our primary hypothesis, we found a robust deficit in the gFc across all diagnostic groups, supporting the notion of a shared cognitive liability that cuts across conventional diagnostic boundaries. Disorder-specific impairments were predominantly observed in NLR particularly among SZ, BD and multiple diagnosis groups, whereas other domains such as CF, WM and SC showed comparatively preserved performance.

Together, these findings reinforce the relevance of a dimensional transdiagnostic framework for understanding cognitive dysfunction in MPI.

In the analyses based on raw test scores, individuals with MPI showed impairment across several cognitive domains, including working memory, verbal learning, cognitive flexibility and social cognition. These deficits were most pronounced in SZ, where impairments were evident in all 4 domains, consistent with the well-established profile of cognitive dysfunction in SZ(43,44). In contrast, individuals with BD, SUD and OCD showed impairment limited to specific domains, suggesting relatively more specific cognitive disruptions in these disorders. However, when we examined latent cognitive constructs - which isolate shared variance across tests, a distinct pattern emerged. A single transdiagnostic impairment in gFc (general factor) was observed across all diagnostic groups supporting the hypothesis that a common cognitive liability underlies multiple psychiatric disorders (10). Because gFc aggregates the shared variance from WM, NLR, CF and SC it reflects the core cognitive dysfunction cutting across diagnoses. Beyond this gFc, only verbal learning deficits remained significant in individuals with SZ, BD and those with multiple diagnosis. This suggests that while gFc accounts for a substantial proportion of transdiagnostic cognitive variability.

A major strength of our work is the use of bi-factor confirmatory factor analytic model to derive neurocognitive indices. Unlike raw test scores or domain averages, which conflate shared cognitive variance with task-specific demands and measurement error, the bifactor structure explicitly separates a global cognitive factor (gFc) from domain-specific components. This distinction is particularly relevant in psychiatric samples, where broad and specific deficits frequently co-occur and may obscure underlying patterns when observed scores are used. By isolating shared variance across cognitive tasks, the model provides a more stable and psychometrically defensible estimate of general cognitive functioning. Consistent with prior work demonstrating that bifactor models improve reliability and interpretability of cognitive constructs, we observed that impairments evident across multiple raw test measures largely converged onto a single latent factor at the structural level(45,46). This supports the notion that cognitive dysfunction across MPIs is driven, to a substantial extent, by a shared cognitive liability rather than multiple independent domain-specific deficits, aligning conceptually with dimensional models of psychopathology such as the general psychopathology (“p”) factor(10).

Beyond the transdiagnostic impairment in gFc, residual domain specific deficits were observed, notably in verbal learning. Even after accounting for shared variance captured by gFc, individuals with SZ, BD and multiple diagnoses continued to show poor performance in verbal learning. This finding suggest that verbal learning may represent a domain which is disproportionately affected in these disorders and not subsumed under gFc. Verbal learning deficits have been consistently reported as prominent features of both SZ and BD(47,48). In contrast, other cognitive domains did not show comparable residual impairments once global cognition was accounted for, underscoring the relative specificity of verbal learning as a distinguishing cognitive domain within the broader transdiagnostic framework.

In examining clinical correlates of cognitive performance, we found that longer duration of illness was modestly but significantly associated with poorer gFc and verbal learning, whereas current illness severity, as indexed by CGI-S was not related to any latent cognitive domain. The association with illness duration suggest that cumulative illness burden may contribute to progressive or sustained cognitive compromise, particularly in gFc and verbal learning(49,50). In contrast, the absence of a relationship with CGI-S should be interpreted in the context of the clinical characteristics of the sample. Participants were largely recruited during relatively stable phases of illness, as reflected by overall low to moderate CGI-S scores across diagnostic groups, a necessary requirement for reliable participation in detailed neuropsychological assessments.

This restricted range of symptom severity likely limited sensitivity to state-dependent cognitive effects. Importantly, this pattern supports the interpretation that the observed transdiagnostic cognitive deficits—especially those captured by gFc—are relatively trait-like and not merely reflections of current symptom burden.

In exploratory analyses of psychotropic exposure, we observed largely limited associations between medication use and latent cognitive performance after adjusting for diagnosis and demographic factors. Antipsychotic use showed an uncorrected association with lower SC scores, but this effect did not survive correction for multiple comparisons, suggesting that medication-related cognitive effects were modest. Antidepressant use was associated with slightly better global cognitive ability, whereas mood stabilisers and benzodiazepines were not associated with any cognitive domain. Given the cross-sectional and observational design, these findings should be interpreted cautiously, as psychotropic prescription is influenced by illness severity and course, raising the possibility of confounding by indication. Overall, the results are consistent with prior evidence(51,52) indicating that cognitive deficits in major psychiatric disorders are only partially modifiable by current pharmacological treatments and represent a core feature of illness rather than a secondary treatment effect.

A key strength of the present study is the examination of cognitive functioning across multiple major psychiatric illnesses within a single, well-characterized cohort using uniform assessment procedures and latent variable modelling. By applying a bifactor confirmatory factor analytic approach and linear mixed-effects models, we were able to distinguish shared and domain-specific cognitive variance while accounting for demographic factors and within-family clustering.

Certain limitations should also be acknowledged. First, cognitive assessment was based on a selected panel of neuropsychological tests rather than a comprehensive battery, reflecting pragmatic constraints related to assessment duration in a large clinical cohort. Second, although demographic factors such as age, sex, and education were accounted for, we were unable to model illness stage or medication dose in a granular manner, given the heterogeneity of illness trajectories and treatment regimens across multiple psychiatric diagnoses. Third, the present findings are based on cross-sectional data and therefore cannot address the longitudinal evolution of cognitive deficits. Ongoing and future analyses within the consortium will examine longitudinal cognitive trajectories in relation to multimodal neuroimaging and genetic data to further elucidate the neurobiological mechanisms underlying transdiagnostic cognitive dysfunction.

## Conclusion

In summary, our findings demonstrate that cognitive dysfunction in MPI is characterized by a robust transdiagnostic impairment in gFc, alongside more limited diagnosis-specific deficits in verbal learning. By applying latent variable modelling within a single, well-characterized cohort, we show that much of the cognitive impairment observed across SZ, BD, SUD, and OCD converges onto a shared cognitive factor that cuts across diagnostic boundaries. These results support dimensional models of psychopathology and highlight gFc as a core feature of MPI. Future longitudinal and mechanistic studies will be essential to clarify the neurobiological substrates and clinical implications of this shared cognitive liability.

## Data Availability

All data produced in the present study are available upon reasonable request to the authors

## Acknowledgements

This research is funded by the Accelerator program for Discovery in Brain disorders using Stem cells (ADBS) (jointly funded by the Department of Biotechnology, Government of India, and the Pratiksha trust; Grant BT/PR17316/MED/31/326/2015), and the Centre for Brain and Mind (CBM) grant of the Rohini Nilekani Philanthropies.

BV is funded by the Intermediate (Clinical and Public Health) Fellowship (IA/CPHI/20/1/505266) of the DBT/Wellcome Trust India Alliance.

A preliminary analysis of this paper with a smaller sample size was published on preprint server (Holla B, Dayal P, Das A, Bhattacharya M, Manjula V, Ithal D, Balachander S, Mahadevan J, Nadella RK, Sreeraj VS, Benegal V. Transdiagnostic neurocognitive endophenotypes in major psychiatric illness. MedRxiv. 2020 Feb 18:2020-02.).

A part of this data was presented as a poster in Bipolar Disorder International Conference (BDICON) 2025 in India (“Can neurocognition differentiate major psychiatric illnesses?”).

## Authors’ Credit statement

Conceptualisation – BV, BH

Methodology – BV, BH

Software – SB, BH

Validation – BV, BH

Statistical analysis – PJ, SB, BH

Investigation – PD, MSJ, AD, MB, AS, CSH, ALS, GJ, MS, SN, AAR, FAS, PV, VT, AK, NJV, RK, CA, TM, SS, APT

Resources – SJ, YCJR

Data curation – PJ, SB, SBN, AS

Writing – Original draft – PJ

Writing – Review & Editing – SB, SBN, BV, BH, UMM, YCJR, SJ

Visualisation – PJ, SB, BH, BV

Supervision – SB, DI, JM, RKN, VSS, GVS, JPJ, VB, YCJR, MV, SJ, BV

Project Administration – VSS, GVS, JPJ, VB, YCJR, MV, SJ, BV Funding Acquisition – SJ, YCJR

## References

1. GBD 2019 Mental Disorders Collaborators. Global, regional, and national burden of 12 mental disorders in 204 countries and territories, 1990-2019: a systematic analysis for the Global Burden of Disease Study 2019. Lancet Psychiatry. 2022 Feb;9(2):137–50.

2. Hyman SE. The diagnosis of mental disorders: the problem of reification. Annu Rev Clin Psychol. 2010;6:155–79.

3. Insel T, Cuthbert B, Garvey M, Heinssen R, Pine DS, Quinn K, et al. Research domain criteria (RDoC): toward a new classification framework for research on mental disorders. Am J Psychiatry. 2010 July;167(7):748–51.

4. Buckley PF, Miller BJ, Lehrer DS, Castle DJ. Psychiatric Comorbidities and Schizophrenia. Schizophr Bull. 2009 Mar;35(2):383–402.

5. Rasic D, Hajek T, Alda M, Uher R. Risk of mental illness in offspring of parents with schizophrenia, bipolar disorder, and major depressive disorder: a meta-analysis of family high-risk studies. Schizophr Bull. 2014 Jan;40(1):28–38.

6. Davies G, Lam M, Harris SE, Trampush JW, Luciano M, Hill WD, et al. Study of 300,486 individuals identifies 148 independent genetic loci influencing general cognitive function. Nat Commun. 2018 May 29;9(1):2098.

7. Brainstorm Consortium, Anttila V, Bulik-Sullivan B, Finucane HK, Walters RK, Bras J, et al. Analysis of shared heritability in common disorders of the brain. Science. 2018 June 22;360(6395):eaap8757.

8. Cross-Disorder Group of the Psychiatric Genomics Consortium, Lee SH, Ripke S, Neale BM, Faraone SV, Purcell SM, et al. Genetic relationship between five psychiatric disorders estimated from genome-wide SNPs. Nat Genet. 2013 Sept;45(9):984–94.

9. Smoller JW, Andreassen OA, Edenberg HJ, Faraone SV, Glatt SJ, Kendler KS. Psychiatric Genetics and the Structure of Psychopathology. Mol Psychiatry. 2019 Mar;24(3):409–20.

10. Caspi A, Houts RM, Belsky DW, Goldman-Mellor SJ, Harrington H, Israel S, et al. The p Factor: One General Psychopathology Factor in the Structure of Psychiatric Disorders? Clin Psychol Sci J Assoc Psychol Sci. 2014 Mar;2(2):119–37.

11. Nieman DH, Chavez-Baldini U, Vulink NC, Smit DJA, van Wingen G, de Koning P, et al. Protocol Across study: longitudinal transdiagnostic cognitive functioning, psychiatric symptoms, and biological parameters in patients with a psychiatric disorder. BMC Psychiatry. 2020 May 11;20(1):212.

12. Abramovitch A, Short T, Schweiger A. The C Factor: Cognitive dysfunction as a transdiagnostic dimension in psychopathology. Clin Psychol Rev. 2021 June;86:102007.

13. Romanowska S, MacQueen G, Goldstein BI, Wang J, Kennedy SH, Bray S, et al. Neurocognitive deficits in a transdiagnostic clinical staging model. Psychiatry Res. 2018 Dec;270:1137–42.

14. Suhas S, Rao NP. Neurocognitive deficits in obsessive–compulsive disorder: A selective review. Indian J Psychiatry. 2019 Jan;61(Suppl 1):S30–6.

15. Millan MJ, Agid Y, Brüne M, Bullmore ET, Carter CS, Clayton NS, et al. Cognitive dysfunction in psychiatric disorders: characteristics, causes and the quest for improved therapy. Nat Rev Drug Discov. 2012 Feb;11(2):141–68.

16. Keramatian K, Torres IJ, Yatham LN. Neurocognitive functioning in bipolar disorder: What we know and what we don’t. Dialogues Clin Neurosci. 2021 Jan 1;23(1):29–38.

17. de Winter L, Jelsma A, Vermeulen JM, van Tricht M, van Weeghel J, Hasson-Ohayon I, et al. Long-Term Changes in Cognition Among Patients With Schizophrenia Spectrum Disorders and Different Durations of Illness: A Meta-Analysis. J Clin Psychiatry. 2024 Sept 25;85(4):23r15134.

18. Husa AP, Moilanen J, Murray GK, Marttila R, Haapea M, Rannikko I, et al. Lifetime antipsychotic medication and cognitive performance in schizophrenia at age 43 years in a general population birth cohort. Psychiatry Res. 2017 Jan;247:130–8.

19. Qi C, Wu Y, Fan Y, Li Y, Wang Z, Zhu H. Effect of mood stabilizers on cognition in bipolar disorder: A systematic review and meta-analysis of randomized controlled trials. J Affect Disord. 2025 Dec 15;391:119982.

20. Viswanath B, Rao NP, Narayanaswamy JC, Sivakumar PT, Kandasamy A, Kesavan M, et al. Discovery biology of neuropsychiatric syndromes (DBNS): a center for integrating clinical medicine and basic science. BMC Psychiatry. 2018 Apr 18;18(1):106.

21. Mehta UM, Thirthalli J, Basavaraju R, Gangadhar BN, Pascual-Leone A. Reduced mirror neuron activity in schizophrenia and its association with theory of mind deficits: evidence from a transcranial magnetic stimulation study. Schizophr Bull. 2014 Sept;40(5):1083–94.

22. Reed GM, Maré KT, First MB, Jaisoorya TS, Rao GN, Dawson-Squibb JJ, et al. The WHO Flexible Interview for ICD-11 (FLII-11). World Psychiatry Off J World Psychiatr Assoc WPA. 2024 Oct;23(3):359–60.

23. Sreeraj VS, Holla B, Ithal D, Nadella RK, Mahadevan J, Balachander S, et al. Psychiatric symptoms and syndromes transcending diagnostic boundaries in Indian multiplex families: The cohort of ADBS study. Psychiatry Res. 2021 Feb;296:113647.

24. D’Elia L, P S, CL U, T W. Color Trails Test (CTT). Psychol Assess Resour. 1996;

25. Lakkireddy SP, Balachander S, Dayalamurthy P, Bhattacharya M, Joseph MS, Kumar P, et al. Neurocognition and its association with adverse childhood experiences and familial risk of mental illness. Prog Neuropsychopharmacol Biol Psychiatry. 2022 Dec 20;119:110620.

26. Owen AM, McMillan KM, Laird AR, Bullmore E. N-back working memory paradigm: a meta-analysis of normative functional neuroimaging studies. Hum Brain Mapp. 2005 May;25(1):46–59.

27. Gopukumar K. RSL. NIMHANS neuropsychology battery 2004 Manual: mannual. Bangalore: NIMHANS; 2004. 267 p. (NIMHANS publicationC; no. 60).

28. Lezak MD, Howieson DB, Bigler ED, Tranel D. Neuropsychological assessment, 5th ed. New York, NY, US: Oxford University Press; 2012. xxv, 1161 p. (Neuropsychological assessment, 5th ed).

29. Valaparla VL, Nehra R, Mehta UM, Thirthalli J, Grover S. Social cognition of patients with schizophrenia across the phases of illness - A longitudinal study. Schizophr Res. 2017 Dec;190:150–9.

30. Perner J, Wimmer H. “John *thinks* that Mary *thinks* that…” attribution of second-order beliefs by 5- to 10-year-old children. J Exp Child Psychol. 1985 June 1;39(3):437–71.

31. Stone VE, Baron-Cohen S, Knight RT. Frontal lobe contributions to theory of mind. J Cogn Neurosci. 1998 Sept;10(5):640–56.

32. Drury VM, Robinson EJ, Birchwood M. “Theory of mind” skills during an acute episode of psychosis and following recovery. Psychol Med. 1998 Sept;28(5):1101–12.

33. Mehta UM, Thirthalli J, Naveen Kumar C, Mahadevaiah M, Rao K, Subbakrishna DK, et al. Validation of Social Cognition Rating Tools in Indian Setting (SOCRATIS): A new test-battery to assess social cognition. Asian J Psychiatry. 2011 Sept 1;4(3):203–9.

34. Busner J, Targum SD. The Clinical Global Impressions Scale. Psychiatry Edgmont. 2007 July;4(7):28–37.

35. Shah AD, Bartlett JW, Carpenter J, Nicholas O, Hemingway H. Comparison of random forest and parametric imputation models for imputing missing data using MICE: a CALIBER study. Am J Epidemiol. 2014 Mar 15;179(6):764–74.

36. Pires L, Moura O, Guerrini C, Buekenhout I, Simões MR, Leitão J. Confirmatory Factor Analysis of Neurocognitive Measures in Healthy Young Adults: The Relation of Executive Functions with Other Neurocognitive Functions. Arch Clin Neuropsychol Off J Natl Acad Neuropsychol. 2019 May 1;34(3):350–65.

37. Becker ML, Ahmed AO, Benning SD, Barchard KA, John SE, Allen DN. Bifactor model of cognition in schizophrenia: Evidence for general and specific abilities. J Psychiatr Res. 2021 Apr;136:132–9.

38. Benjamini Y, Hochberg Y. Controlling the False Discovery Rate: A Practical and Powerful Approach to Multiple Testing. J R Stat Soc Ser B Methodol. 1995;57(1):289–300.

39. Buuren S van, Groothuis-Oudshoorn K. mice: Multivariate Imputation by Chained Equations in R. J Stat Softw. 2011 Dec 12;45:1–67.

40. Rosseel Y. lavaan: An R Package for Structural Equation Modeling. J Stat Softw. 2012 May 24;48:1–36.

41. Bates D, Mächler M, Bolker B, Walker S. Fitting Linear Mixed-Effects Models Using lme4. J Stat Softw. 2015 Oct 7;67:1–48.

42. Hu L, Bentler PM. Cutoff criteria for fit indexes in covariance structure analysis: Conventional criteria versus new alternatives. Struct Equ Model Multidiscip J. 1999 Jan 1;6(1):1–55.

43. Nuechterlein KH, Nasrallah H, Velligan D. Measuring Cognitive Impairments Associated With Schizophrenia in Clinical Practice: Overview of Current Challenges and Future Opportunities. Schizophr Bull. 2025 Mar 1;51(2):401–21.

44. McCutcheon RA, Keefe RSE, McGuire PK. Cognitive impairment in schizophrenia: aetiology, pathophysiology, and treatment. Mol Psychiatry. 2023 May;28(5):1902–18.

45. Gignac GE, Watkins MW. Bifactor Modeling and the Estimation of Model-Based Reliability in the WAIS-IV. Multivar Behav Res. 2013 Sept;48(5):639–62.

46. Reise SP. The Rediscovery of Bifactor Measurement Models. Multivar Behav Res. 2012 Sept 1;47(5):667–96.

47. Robinson LJ, Thompson JM, Gallagher P, Goswami U, Young AH, Ferrier IN, et al. A meta-analysis of cognitive deficits in euthymic patients with bipolar disorder. J Affect Disord. 2006 July;93(1–3):105–15.

48. Kahn RS, Keefe RSE. Schizophrenia is a cognitive illness: time for a change in focus. JAMA Psychiatry. 2013 Oct;70(10):1107–12.

49. Bowie CR, Harvey PD. Cognition in schizophrenia: impairments, determinants, and functional importance. Psychiatr Clin North Am. 2005 Sept;28(3):613–33, 626.

50. Lewandowski KE, Cohen BM, Ongur D. Evolution of neuropsychological dysfunction during the course of schizophrenia and bipolar disorder. Psychol Med. 2011 Feb;41(2):225–41.

51. Keefe RSE, Bilder RM, Davis SM, Harvey PD, Palmer BW, Gold JM, et al. Neurocognitive effects of antipsychotic medications in patients with chronic schizophrenia in the CATIE Trial. Arch Gen Psychiatry. 2007 June;64(6):633–47.

52. Millan MJ, Agid Y, Brüne M, Bullmore ET, Carter CS, Clayton NS, et al. Cognitive dysfunction in psychiatric disorders: characteristics, causes and the quest for improved therapy. Nat Rev Drug Discov. 2012 Feb 1;11(2):141–68.

