## Supplemental Table S1 for "A unified general cognitive factor explains impairment across schizophrenia, bipolar disorder, OCD and substance use disorder"

*Table S1 Combination of diagnosis in multiple diagnosis group*

| Diagnosis | Numbers (Total n = 72) |
| --- | --- |
| SZ + SUD | 20 |
| BD +SUD | 19 |
| SZ + OCD | 19 |
| Schizoaffective disorder | 7 |
| BD + OCD | 4 |
| OCD + SUD | 3 |

SZ – Schizophrenia, BD – Bipolar disorder, SUD – substance use disorder, OCD – obsessive compulsive disorder
