## Supplemental Table S2 for "A unified general cognitive factor explains impairment across schizophrenia, bipolar disorder, OCD and substance use disorder"

*Table S2 Effects of diagnosis on neurocognition (raw scores) using linear mixed effects analysis covarying age, gender and years of education (expressed as Beta and* [*C.Is*](http://c.is)*), *p value < 0.05, ** FDR adjusted p < 0.05*

| **Predictor** | **ETC1** | **ETC2** | **NB1_Acc** | **NB2_Acc** | **AVLT_IR** | **AVLT_TOTAL** | **SOT** |
| --- | --- | --- | --- | --- | --- | --- | --- |
| Predictors of interest | | | | | | | |
| BD | -0.07 [-0.29, 0.15] | -0.29 [-0.51, -0.08]* | -0.38 [-0.59, -0.18]** | -0.38 [-0.58, -0.17]** | -0.6 [-0.8, -0.41]** | -0.7 [-0.88, -0.51]** | -0.16 [-0.39, 0.06] |
| Multiple | -0.13 [-0.39, 0.13] | -0.11 [-0.36, 0.15] | -0.49 [-0.74, -0.25]** | -0.53 [-0.77, -0.29]** | -0.73 [-0.95, -0.5]** | -0.81 [-1.02, -0.6]** | -0.25 [-0.51, 0] |
| OCD | -0.06 [-0.27, 0.15] | -0.1 [-0.3, 0.11] | -0.16 [-0.36, 0.03] | -0.21 [-0.41, -0.02]* | -0.16 [-0.35, 0.03] | -0.22 [-0.39, -0.04]* | -0.24 [-0.45, -0.03]* |
| SUD | -0.29 [-0.51, -0.07]* | -0.13 [-0.35, 0.1] | -0.32 [-0.53, -0.11]** | -0.44 [-0.65, -0.23]** | -0.38 [-0.58, -0.18]** | -0.39 [-0.58, -0.2]** | -0.17 [-0.39, 0.06] |
| SZ | -0.25 [-0.48, -0.02]* | -0.42 [-0.66, -0.19]** | -0.61 [-0.83, -0.39]** | -0.5 [-0.72, -0.28]** | -0.79 [-1, -0.58]** | -0.93 [-1.13, -0.73]** | -0.44 [-0.67, -0.2]** |
| Covariates | | | | | | | |
| Age | -0.09 [-0.16, -0.02]* | -0.1 [-0.17, -0.03]** | -0.09 [-0.15, -0.02]* | -0.13 [-0.19, -0.06]** | -0.25 [-0.3, -0.19]** | -0.26 [-0.32, -0.21]** | -0.06 [-0.13, 0] |
| Gender | 0.16 [0.01, 0.3]* | -0.03 [-0.18, 0.11] | 0.19 [0.06, 0.33]** | 0.18 [0.05, 0.32]* | -0.24 [-0.37, -0.12]** | -0.28 [-0.4, -0.16]** | -0.16 [-0.31, -0.02]* |
| Years of education | 0.23 [0.16, 0.31]** | 0.24 [0.16, 0.31]** | 0.27 [0.2, 0.35]** | 0.29 [0.22, 0.36]** | 0.24 [0.17, 0.31]** | 0.26 [0.2, 0.33]** | 0.2 [0.12, 0.27]** |
