## Supplemental Table S3 for "A unified general cognitive factor explains impairment across schizophrenia, bipolar disorder, OCD and substance use disorder"

Supplementary Table S3 Spearmann correlations between latent cognitive domains and clinical severity and chronicity variables *p value < 0.05

| Latent cognitive domain | CGI – S ρ (p value) | TDI ρ (p value) |
| --- | --- | --- |
| gFc | 0.05 (0.21) | -0.19 (< 0.001)* |
| NLR | 0.05 (0.24) | -0.17 (< 0.001)* |
| WM | 0.01 (0.70) | 0.04 (0.45) |
| CF | -0.05 (0.23) | -0.05 (0.32) |
| SC | -0.03 (0.45) | -0.03 (0.42) |
