## Supplemental Table S4 for "A unified general cognitive factor explains impairment across schizophrenia, bipolar disorder, OCD and substance use disorder"

TableS4 Effect of diagnosis and treatment variables on latent domains of neurocognition using linear mixed effects analysis covarying for age, gender and years of education * p < 0.05, ** FDR adjusted p < 0.05

| Fixed effects | gFc | NLR | WM | CF | SC |
| --- | --- | --- | --- | --- | --- |
| Predictors of Interest: Diagnoses and Treatment variables | | | | | |
| BD | -0.66 [-1.01,-0.32]** | -0.24 [-0.63,0.14] | -0.11 [-0.40,0.18] | -0.30 [-0.78, 0.18] | 0.00 [-0.37, 0.38] |
| Multiple diagnosis | -0.87 [-1.17,-0.57]** | -0.45 [-0.79,-0.11]** | -0.22 [-0.47,0.04] | -0.18 [-0.60,0.24] | 0.13 [-0.20,  0.46] |
| OCD | -0.42 [-0.70,-0.15]** | -0.06 [-0.37,0.25] | -0.05 [-0.29,0.18] | -0.23 [-0.62,0.15] | -0.24 [-0.55, 0.06] |
| SUD | -0.70 [-0.91,-0.49]** | -0.21 [-0.45,0.03] | -0.09 [-0.27,0.09] | -0.42 [-0.72,-0.13]** | -0.19 [-0.43, 0.04] |
| SZ | -0.96 [-1.23,-0.68]** | -0.53 [-0.85,-0.22]** | -0.12 [-0.35,0.12] | -0.16 [-0.55,0.23] | 0.01 [-0.30, 0.31] |
| Antipsychotic (Yes) | 0.12 [-0.07,0.31] | 0.01 [-0.00,0.03] | 0.00 [-0.17,0.17] | 0.14 [-0.14,0.41] | -0.25 [-0.46, -0.03]* |
| Mood Stabiliser  (Yes) | 0.10 [-0.16,0.36] | 0.01 [-0.21,0.23] | 0.05 [-0.17,0.28] | 0.29 [-0.08, 0.65] | 0.17 [-0.11, 0.46] |
| Antidepressant  (Yes) | 0.24 [0.02,0.45]** | 0.03 [-0.21,0.27] | -0.02 [-0.20,0.16] | 0.19 [-0.11,0.49] | 0.19 [-0.05, 0.42] |
| Benzodiazepine  (Yes) | -0.07 [-0.46,0.32] | -0.02 [-0.46,0.42] | -0.17 [-0.50,0.17] | 0.16 [-0.39,0.70] | 0.24 [-0.19, 0.67] |
| Covariates | | | | | |
| Age | -0.01  [-0.02,-0.01]** | -0.02  [-0.02,-0.01]** | 0.001  [-0.00,0.00] | -0.001  [-0.01,0.00] | -0.002  [-0.00,0.00] |
| Gender (Male) | 0.02  [-0.08,0.13] | -0.34  [-0.46,-0.23]** | 0.14  [0.04,0.24]** | 0.17  [0.01,0.33]* | -0.10  [-0.23,-0.02] |
| Years of education | 0.07  [0.06,0.08]** | 0.01  [-0.00,0.03] | 0.01  [0.00,0.03]** | 0.02  [0.01,0.04]* | 0.02  [0.00,0.03]* |
